# Low grade systemic inflammation is a risk factor for infection death: an observational cohort study

**DOI:** 10.1101/2021.11.08.21266043

**Authors:** Michael Drozd, Mar Pujades-Rodriguez, Ann W Morgan, Patrick J Lillie, Klaus K Witte, Mark T Kearney, Richard M Cubbon

## Abstract

**Background:** Many diseases are associated with chronic inflammation, resulting in widening indications for anti-inflammatory therapies. Whilst effective as disease modifying agents, these increase the risk of serious infection.

**Objective:** To determine if low-grade inflammation is associated with fatal infection, irrespective of associated comorbidity or anti-inflammatory therapy.

**Design:** Observational cohort study

**Setting:** UK Biobank study

**Participants:** 461,052 people

**Interventions:** None

**Measurements:** Incidence rate ratio (IRR) of death from infection, cardiovascular disease, or other causes, adjusted for comorbidities and use of anti-inflammatory therapies, for serum C-reactive protein (CRP) at recruitment.

**Results:** Low grade inflammation was common in all morbidities considered and was more prevalent as multimorbidity accrued (CRP ≥2mg/L in 23.3% of people without disease and 58.7% with 3+ comorbidities; p<0.001). After adjusting for confounding factors, CRP ≥2mg/L was associated with a higher IRR of infection death (IRR 1.70; 95% confidence interval 1.51-1.92) than cardiovascular death (IRR 1.48; 1.40-1.57) or other causes of death (IRR 1.41; 1.37-1.45); CRP thresholds of ≥5 and ≥10 mg/L yielded similar findings. Absolute rates of infection, cardiovascular and other death were 0.43, 1.59 and 5.39 per 1000 participant-years, respectively, in people with CRP ≥2mg/L. Analyses stratified by disease type, or number of comorbidities, showed consistent associations between elevated CRP and infection death.

**Limitations:** Our observational study design precludes assessment of causality. We lacked data on the use of anti-inflammatory therapies after study recruitment.

**Conclusion:** Low grade inflammation, irrespective of associated comorbidity, identifies people at particularly increased risk of infection death. Decisions to use anti-inflammatory therapies guided by low grade inflammation require careful consideration of the associated risks and benefits.

---

Inflammation is a common pathological factor in many chronic diseases including atherosclerosis, arthritis, chronic lung disease, cancer, diabetes and obesity;(1–5) moreover, it is more common as multimorbidity accrues.(6–8) The success of anti-inflammatory therapies as disease modifying agents for inflammatory rheumatological, dermatological and gastrointestinal disorders has recently prompted phase 3 clinical trials in the context of atherosclerosis.(9,10) There is also hope that inflammation could represent a novel therapeutic target in diseases ranging from heart failure to cancer to depression.(11–13) However, canakinumab and colchicine failed to improve overall survival in people with advanced atherosclerosis, in spite of substantially reducing cardiovascular events, probably because of the increased risk of fatal infections.(9,14) This highlights the complexity of therapeutic modulation and suggests that future approaches will require nuance,(15) perhaps informed by experience from more established indications for anti-inflammatory therapy.(16) However, in spite of a wealth of experience in therapeutically targeting inflammation, it remains unclear whether systemic inflammation *per se* is a risk factor for serious infection, possibly as a biomarker of more broadly perturbed immune responses to pathogens. We set out to answer this question using the UK Biobank (UKB) cohort study, which provides detailed phenotyping of approximately 500,000 adults, including large numbers with diverse chronic inflammatory morbidities.

## Methods

### UK Biobank cohort

UKB is a population-based prospective cohort study that consists of 502,505 people aged between 37-73 years. The resource was developed using UK Government and biomedical research charity funding to improve understanding of disease and is an open access resource for all *bona fide* researchers. Full details of the study design and conduct are available from the UKB website (https://www.ukbiobank.ac.uk). Participants were recruited between 2006 and 2010, and attended one of 22 assessment centres across England, Scotland and Wales. Whilst the cohort is not representative of the whole UK population relating to socio-economic deprivation (SED), some non-communicable diseases and ethnic minorities, it allows assessment of exposure-disease relationships.(17) Baseline biological measurements were recorded, and participants completed a touchscreen and nurse-led questionnaire, as described elsewhere.(18) UKB received ethical approval from the NHS Research Ethics Service (11/NW/0382); we conducted this analysis under application number 59585. All participants provided written informed consent.

### Definitions of systemic inflammation and study covariates

Low-grade systemic inflammation was defined using serum C-reactive protein (CRP) data generated with a high sensitivity immunoturbidimetric assay (Beckman Coulter AU5800). UKB collected CRP data at study enrolment from 468,528 participants. Our primary analyses defined systemic inflammation as CRP ≥2mg/L, based upon prior clinical trials targeting anti-inflammatory agents to people above this threshold.(9,19) Potential confounding factors were age, sex, ethnicity, SED, smoking status, comorbidity and anti-inflammatory medical therapy, all determined at study recruitment. Ethnicity was participant-classified within UKB-defined categories of white, mixed, Asian or British Asian, black or British black, Chinese or other ethnic group; due to the small number of people (and deaths) in each minority (non-white) ethnic group, these were pooled as ‘non-white ethnicity’. Smoking status was self-reported as never, previous or current at the point of recruitment. SED was measured using the Townsend score, an area-based deprivation index, and categorised into quintiles. Obesity was classified using the World Health Organisation’s categorisation according to body mass index: class 1 (30.0-34.9 kg/m^2^), class 2 (35.0-39.9 kg/m^2^), class 3 (≥40 kg/m^2^). Self-reported medical disorders recorded solely at study recruitment during face-to-face interview with a nurse were used to classify morbidities (described in **Appendix Table 1**). We used clinical consensus prior to our analyses to select a range of morbidities that represent a broad spectrum of common disease groups: hypertension, chronic heart disease (ischaemic heart disease and heart failure), chronic respiratory disease, diabetes, prior cancer, chronic liver disease, chronic kidney disease, prior stroke or transient ischaemic attack (TIA), other neurological disease, psychiatric disease and chronic inflammatory and autoimmune rheumatic disease.(20) The number of these morbidities was calculated for each participant as an index of multimorbidity. Self-reported use of non-steroidal anti-inflammatory or immunosuppressive agents (including disease-modifying anti-rheumatic drugs and oral glucocorticoids) was assessed at study enrolment as described in **Appendix Table 2**. Missing data for comorbidities (n=863), body mass index (n=3,106), smoking (n=2,949), ethnicity (n=2,777), and SED (n=624), and loss to follow□up or withdrawal of consent (n=1,343), resulted in exclusion of 7,476 participants from our analyses (some participants with more than one variable missing), resulting in a study cohort of 461,052 participants.

### Definition of outcomes

Mortality information provided by UKB was derived from linked national death registry data from NHS Digital for participants in England & Wales and from the NHS Central Register, part of the National Records of Scotland, for participants in Scotland. In the present analysis, we censored follow-up and only considered deaths until 31^st^ December 2019 to ensure this was before the first reported case of COVID-19 in the United Kingdom.(21) As we have previously described, deaths were classified using ICD-10 codes for the main cause of death as infection-related,(20) cardiovascular,(22) or other causes; specific codes are described in **Appendix Table 3**. Infection death was our primary study outcome. Cardiovascular death was a secondary outcome given the wealth of data causally linking low-grade systemic inflammation to adverse cardiovascular outcomes.(1,9,10,15)

### Statistical analysis

Continuous variables are presented as mean (standard deviation) or median (inter-quartile range) if non-normally distributed, and categorical variables as number (percentage). Adjusted cause-specific mortality incidence rate ratios (IRR) were estimated using Poisson regression with exposure time modelled. Time-varying covariates were not used, and the calendar year of recruitment was not included in models due to the narrow recruitment era. Unless specified otherwise, models were adjusted for all of the following covariates: age, sex, SED, smoking status, obesity, hypertension, chronic heart disease, chronic respiratory disease, diabetes, cancer, chronic liver disease, chronic kidney disease, prior stroke/TIA, other neurological disease, psychiatric disorder, autoimmune rheumatological disease, NSAID and immunosuppressive agent use. CRP was dichotomised as 2 or ≥2 mg/L in our primary analyses since this threshold has been applied in clinical trials of anti-inflammatory therapy.(9,14) When addressing multimorbidity, we separately modelled the number of comorbidities (1, 2, and 3 or more, since few participants had 4 or more) in place of the individual comorbidity variables (obesity, hypertension, chronic heart disease, chronic respiratory disease, diabetes, cancer, chronic liver disease, chronic kidney disease, prior stroke/TIA, other neurological disease, psychiatric disorder and rheumatological disease). When stratifying the population by specific morbidities, or the number of comorbidities, the stratifying factor was excluded from the model. Correlation matrices of Poisson model’s coefficients were used to assess correlation between covariates; no correlation coefficients >0.3 or <-0.3 were observed. As previously described,(20) age was modelled using restricted cubic splines with 5 knots for infection death analyses and 4 knots for non-infection death analyses, since these provided the best fit as assessed by the Akaike and the Bayesian Criterion (models including categorical, linear, cubic splines with 3, 4 and 5 knots and first and second degree fractional polynomials were compared). Secondary analyses included: 1) assessment of age/sex adjusted, age/sex/socio-demographic factor/comorbidity adjusted and fully adjusted models; 2) sub-group analyses stratified by specific disease states or multimorbidity categories. Sensitivity analyses included: 1) assessment of alternate CRP thresholds of ≥5mg/L and ≥10mg/L; and 2) exclusion of participants who died during the first 6 months of follow-up, since elevated CRP could denote acute infection. All tests were 2-sided and statistical significance was defined as p<0.05. All statistical analyses were performed using Stata/MP (StataCorp LP, College Station, USA; version 16.1).

## Results

Within a study population of 461,052 people, 35.2% (n=162,419) had serum CRP ≥2mg/L (characteristics of participants without CRP data are presented in **Appendix Table 4**). In comparison to people with CRP <2mg/L, those with CRP ≥2mg/L were older, and more frequently female, socio-economically deprived, current smokers, and were more likely to have a range of chronic illnesses (**Table 1**). Similar observations resulted from analyses of people with CRP ≥5mg/L (11.6%; n=53,468) and ≥10mg/L (4.1%; n=19,024), as shown in **Appendix Tables 5-6**. Notably, CRP ≥2mg/L was highly prevalent in all of the chronic diseases studied, ranging from 39.6% of people with cancer, to 85.6% of people with class 3 obesity, and was more prevalent in people with greater multimorbidity (**Figure 1**).

**Table 1:**
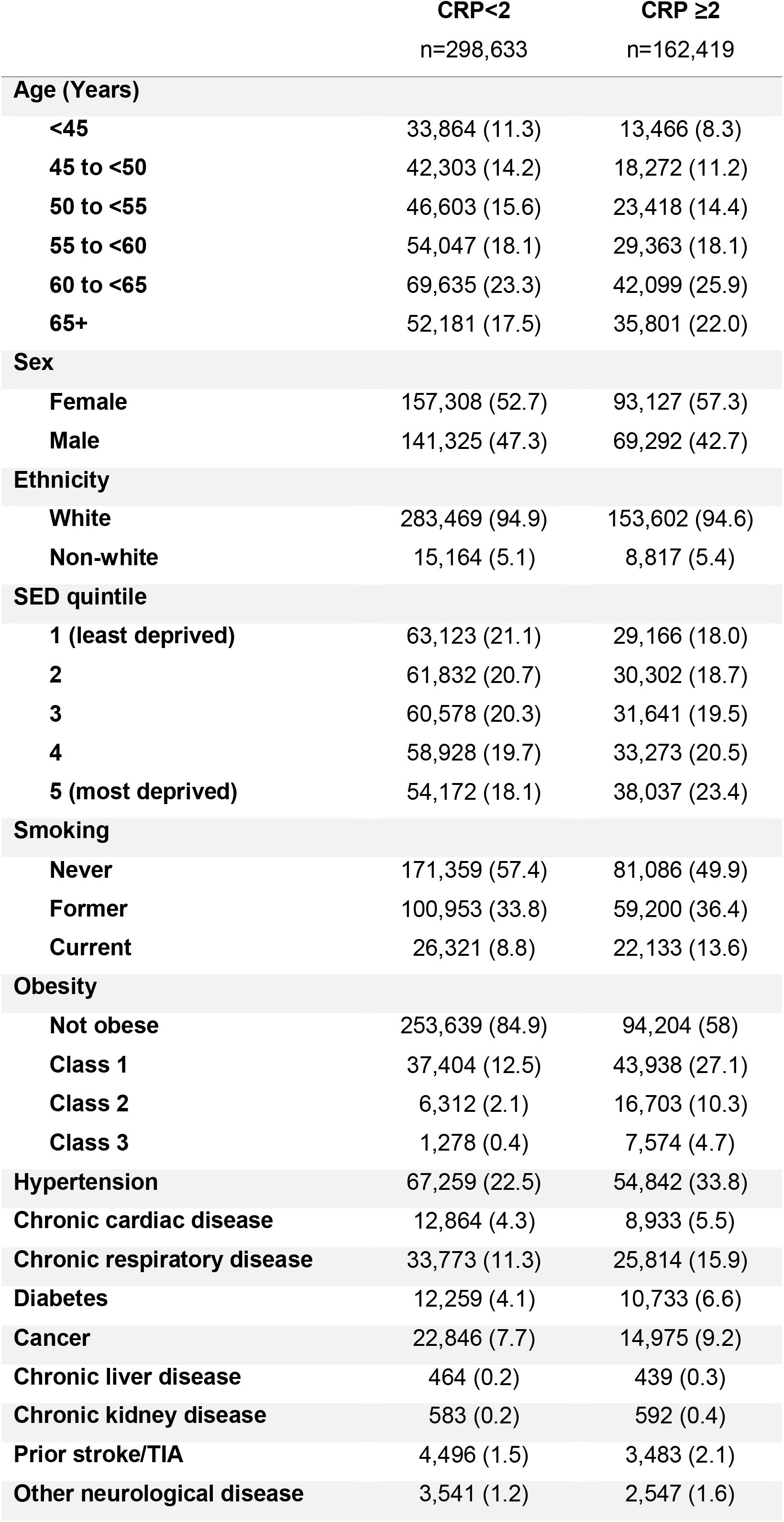
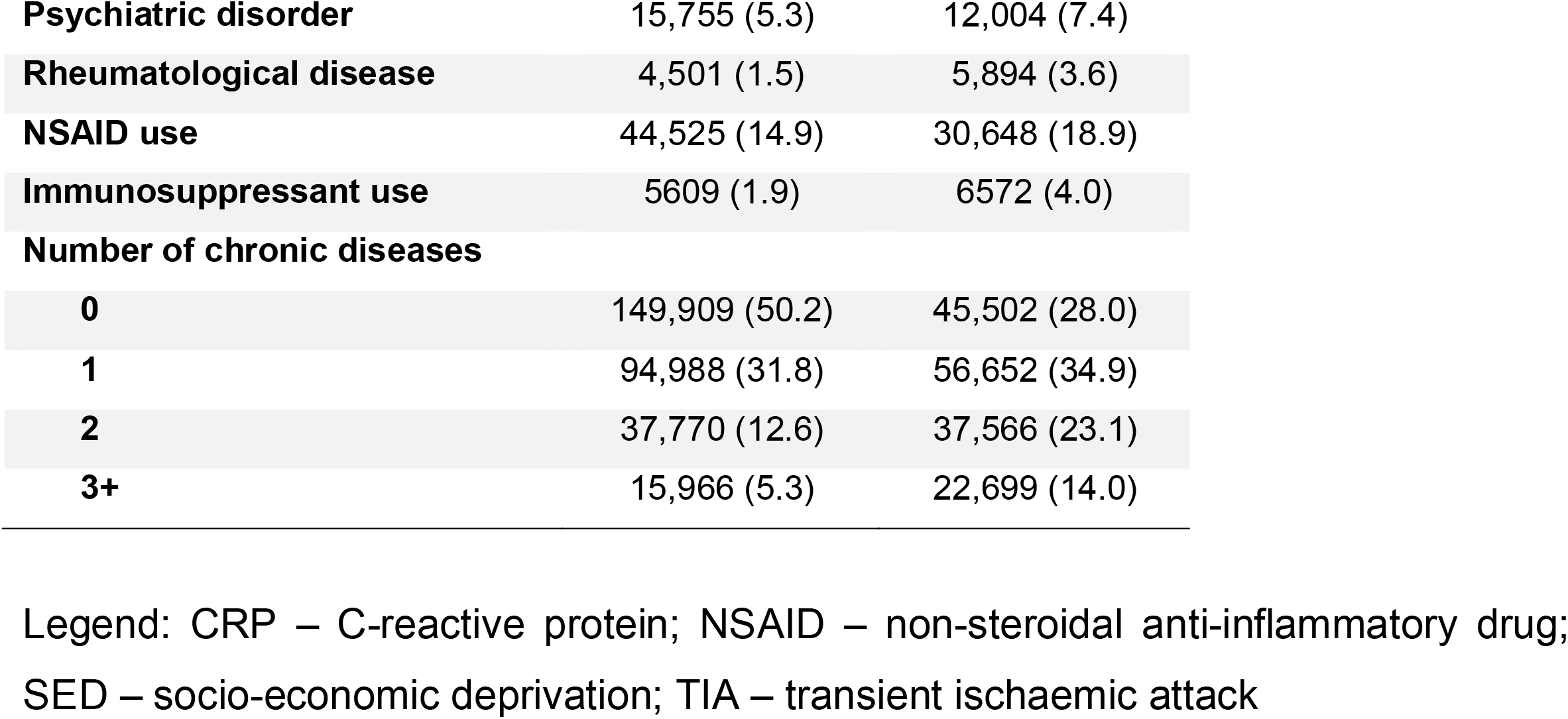
Characteristics of people with CRP <2mg/L and ≥2mg/L.

**Figure 1:**
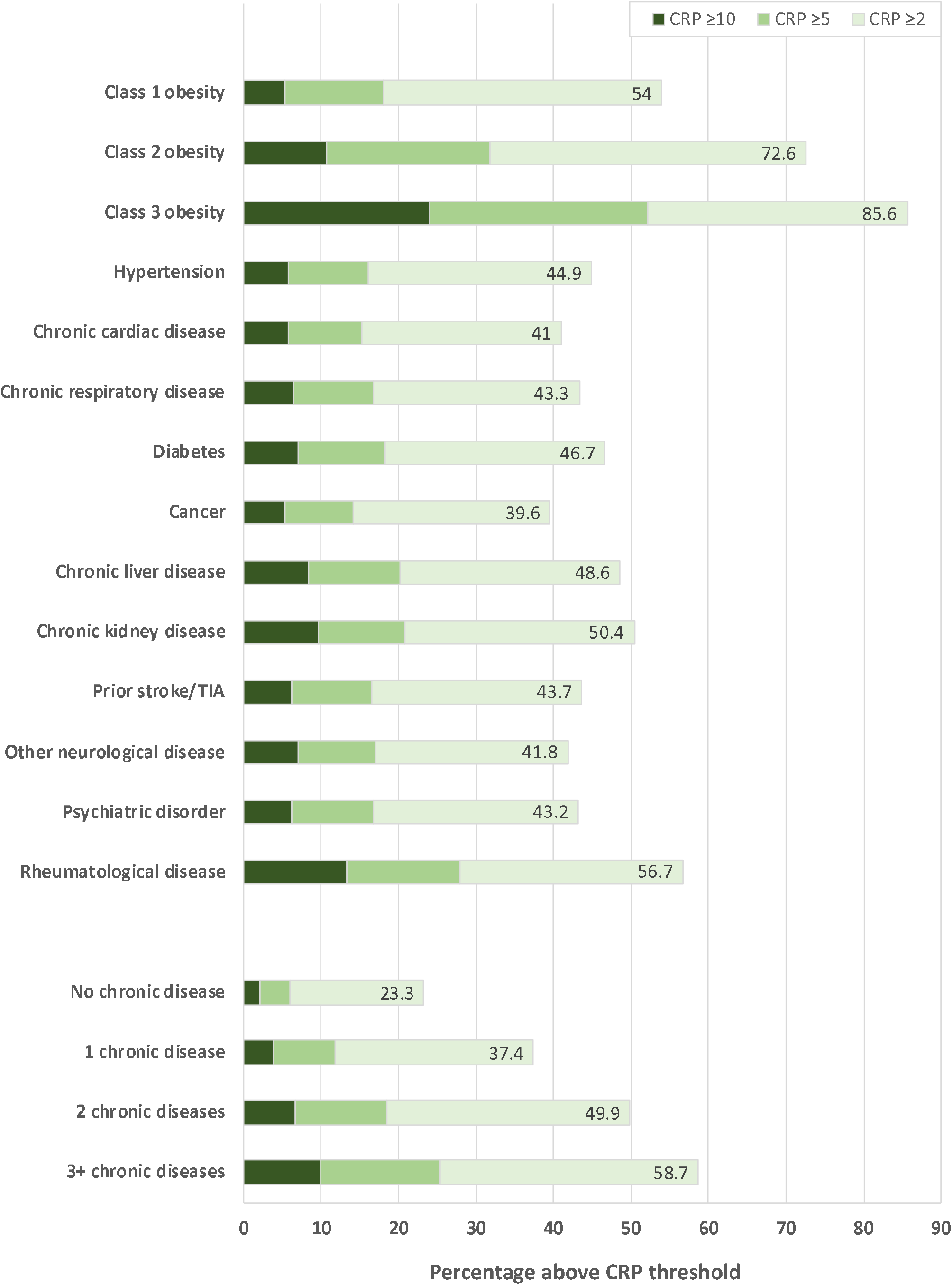
Prevalence of elevated CRP in chronic morbidity or multimorbidity groups. Stacked bar chart illustrating the percentage of people in specified chronic disease and multimorbidity groups with CRP ≥2mg/L, ≥5mg/L and ≥10mg/L. CRP – C-reactive protein; TIA – transient ischaemic attack.

After 4,927,012 person-years of follow-up (median 10.9 [IQR 10.1 – 11.6] years per participant), 25,619 deaths (5.6% of participants) occurred. Of these, 1,274 (5.0%) were attributed to infection, 5,202 (20.3%) to cardiovascular events, and 19,143 (74.7%) to other causes. IRRs for the association between CRP ≥2mg/L and these three categories of death are shown in **Figure 2**. Results from secondary analyses presenting IRRs from unadjusted, age/sex adjusted, and other models are also presented in **Appendix Table 7**. To exclude the possibility that deaths occurring early during follow-up were related to undiagnosed acute infection at the time of enrolment, we repeated analyses after excluding all deaths during the first 6-months and derived similar IRRs (**Appendix Table 8**). These data illustrate that CRP ≥2mg/L is associated with increased risk of the three categories of death, but with the relative risk of infection death being consistently higher than for cardiovascular or other death. Sensitivity analyses using higher CRP thresholds of ≥5mg/L and ≥10mg/L yield the same conclusion (**Figure 2)**. However, in spite of higher relative risk of infection death, it is important to emphasise that the absolute rates of infection death was lower than those of cardiovascular and other death, even in people with CRP ≥10mg/L (**Table 2**).

**Table 2:** Absolute cause-specific mortality rates according to CRP category.

|  | Rates per 1000 person years (95% CI) |  |  |
| --- | --- | --- | --- |
|  | Infection death | CV death | Other death |
| **CRP $\geq 2$** | 0.43 (0.40-0.46) | 1.59 (1.53-1.65) | 5.39 (5.28-5.50) |
| **CRP $\geq 5$** | 0.64 (0.58-0.71) | 2.08 (1.96-2.20) | 6.83 (6.62-7.05) |
| **CRP $\geq 10$** | 0.88 (0.76-1.02) | 2.41 (2.20-2.64) | 8.53 (8.14-8.95) |

Legend: CI – confidence interval; CRP – C-reactive protein; CV – cardiovascular

**Figure 2:**
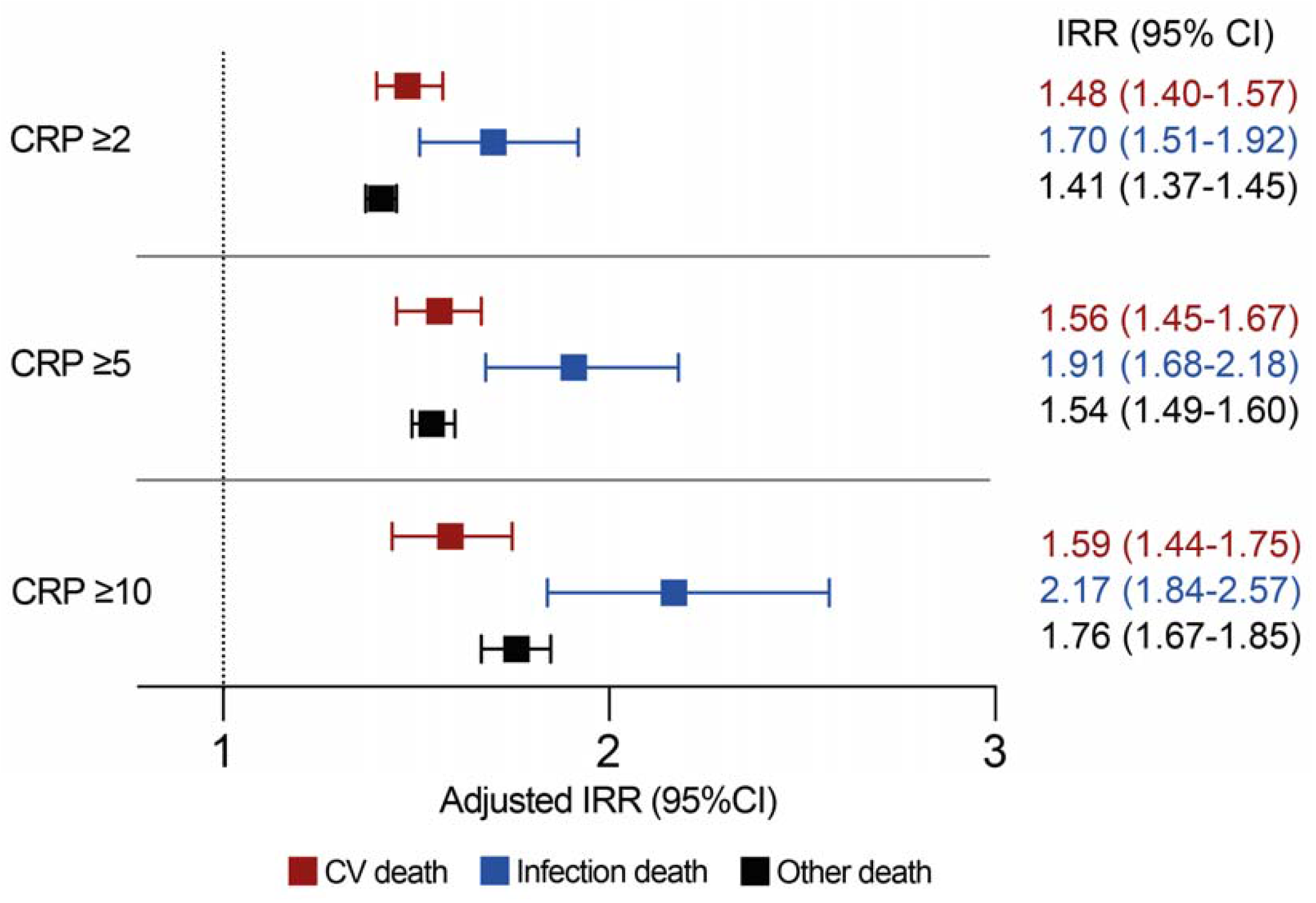
Adjusted incidence rate ratios of cause-specific mortality according to CRP category. Forest plot illustrating adjusted incidence rate ratios (IRR) and their 95% confidence intervals (CI) for specified modes of death in people with CRP ≥2mg/L, ≥5mg/L and ≥10mg/L (versus below these thresholds). The adjusted model includes the following factors in addition to CRP categories: age, sex, SED, smoking status, obesity, hypertension, chronic heart disease, chronic respiratory disease, diabetes, cancer, chronic liver disease, chronic kidney disease, prior stroke/TIA, other neurological disease, psychiatric disorder, autoimmune rheumatological disease, self-reported NSAID prescription and self-reported immunosuppressive agent prescription. CV – cardiovascular.

We also assessed whether the association between low-grade systemic inflammation and cause-specific mortality was consistent in subgroups with specific morbidities or accumulating multimorbidity. Again, CRP ≥2mg/L was associated with increased risk of all categories of death, but nominally higher IRRs were observed for infection death than cardiovascular or other death in all morbidities except ‘other neurological diseases’ (**Figure 3**). The same conclusion was reached irrespective of the number of comorbid diseases, although the IRR for infection death was nominally lower than that for cardiovascular death in people without disease (**Figure 4**). Sensitivity analyses applying higher CRP thresholds of ≥5mg/L and ≥10mg/L yielded similar conclusions (**Appendix Tables 9-12**).

**Figure 3:**
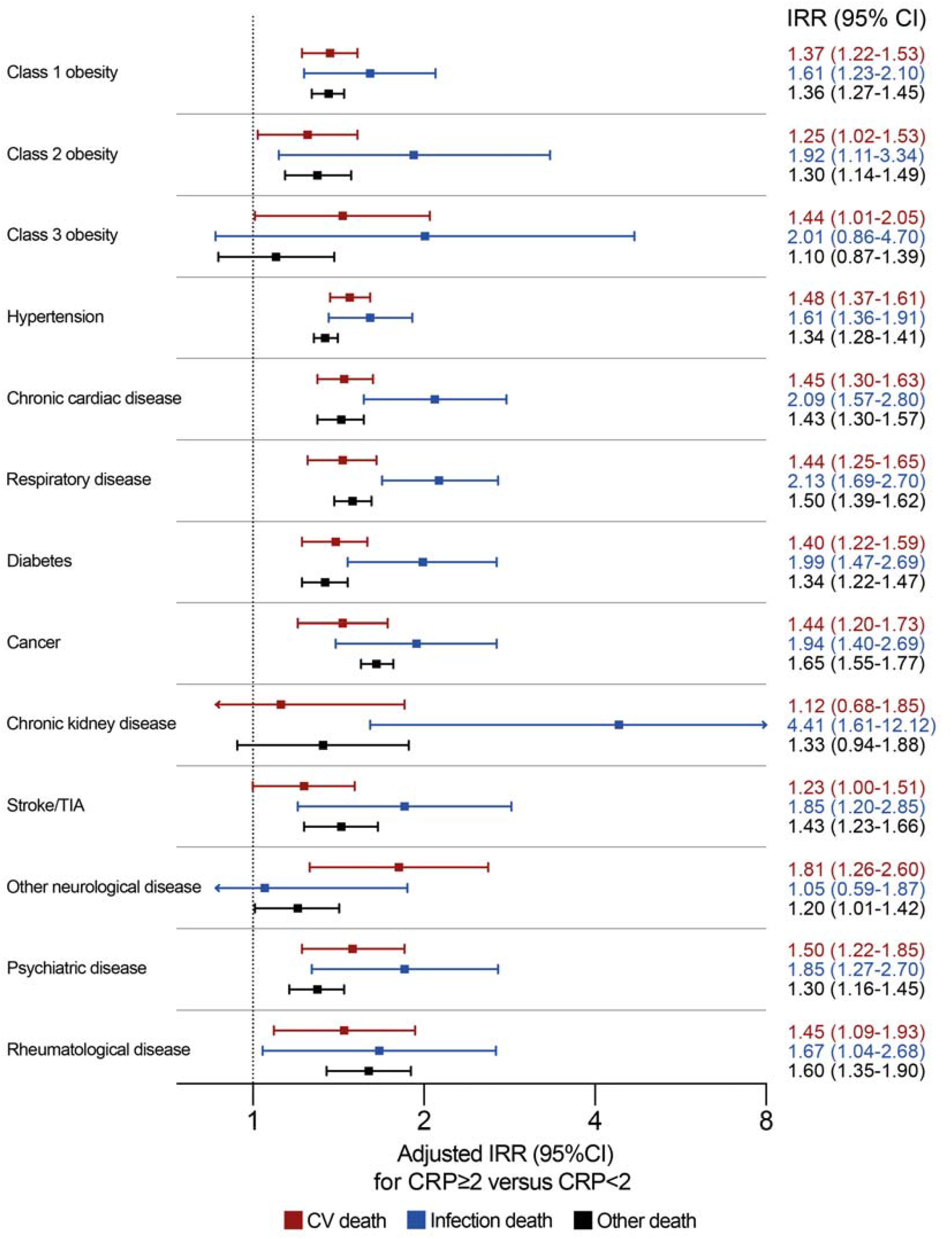
Adjusted incidence rate ratios of cause-specific mortality in people with CRP ≥2mg/L stratified by chronic disease group. Forest plot illustrating adjusted incidence rate ratios (IRR) and their 95% confidence intervals (CI) for specified modes of death in people with CRP ≥2mg/L versus CRP <2mgL stratified by chronic disease group. The adjusted model includes the following factors in addition to CRP status within chronic disease group strata: age, sex, SED, smoking status, comorbidity beyond defined strata (including obesity, hypertension, chronic heart disease, chronic respiratory disease, diabetes, cancer, chronic liver disease, chronic kidney disease, prior stroke/TIA, other neurological disease, psychiatric disorder) autoimmune rheumatological disease), self-reported NSAID prescription and self-reported immunosuppressive agent prescription. CV – cardiovascular; TIA – transient ischaemic attack.

**Figure 4:**
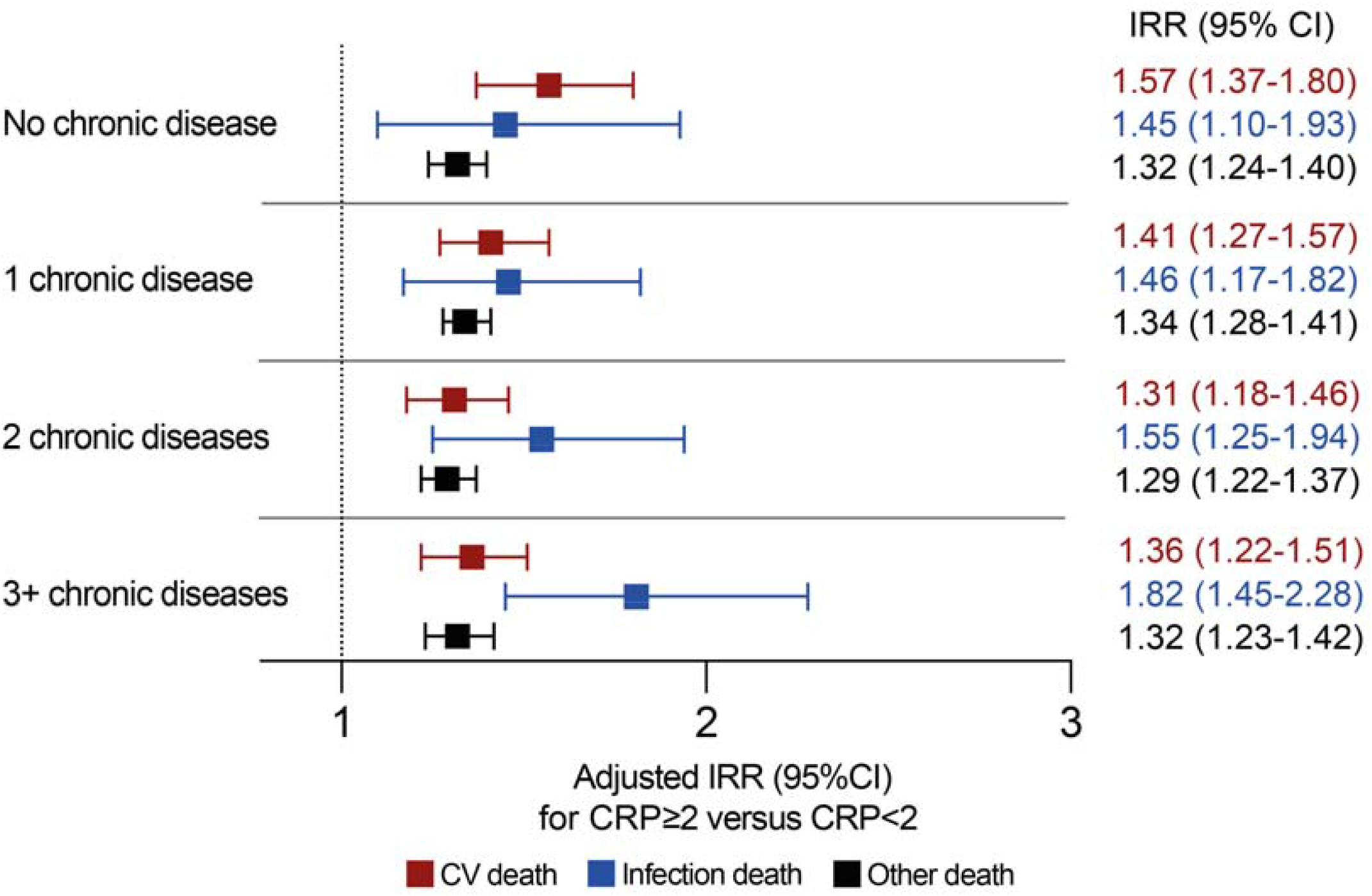
Adjusted incidence rate ratios of cause-specific mortality in people with CRP ≥2mg/L stratified by number of morbidities. Forest plot illustrating adjusted incidence rate ratios (IRR) and their 95% confidence intervals (CI) for specified modes of death in people with CRP ≥2mg/L versus CRP <2mgL stratified by the extent of multimorbidity. The adjusted model includes the following factors in addition to CRP status within multimorbidity strata: age, sex, SED, smoking status, self-reported NSAID prescription and self-reported immunosuppressive agent prescription. CV – cardiovascular.

## Discussion

Our analysis is the first to explore the association between low-grade systemic inflammation and infection death and it has important implications for future research and clinical practice. We show that elevated CRP is highly prevalent in many morbidities and is associated with a greater relative risk of infection death than cardiovascular or other causes of death, irrespective of the CRP threshold chosen. This observation was consistent in stratified analyses across the vast majority of diseases we studied, suggesting that it is broadly relevant to people with diverse diseases, or combinations of diseases. In the context of broadening clinical use of anti-inflammatory therapies, our data caution that people with elevated CRP are inherently predisposed to fatal infection. This suggests that careful balancing of risks and benefits of such therapies is essential, which is likely to require greater understanding of how perturbed inflammation contributes to chronic diseases in order to personalise therapy.

Our findings may be particularly pertinent to recent clinical trials, which targeted canakinumab or colchicine to people with advanced atherosclerotic cardiovascular disease.(9,10) Inflammation is a key driver of atherosclerosis,(1,15) and these trials demonstrated impressive reductions in adverse cardiovascular events, yet without improving overall mortality.(9,10) Serious infection events are were also substantially increased by these agents,(9,14) which may underpin their failure to improve overall survival in people with advanced atherosclerosis; this represents a major hurdle to routine clinical adoption. Whilst these trials only recruited people after myocardial infarction, data from people with other cardiovascular diseases demonstrate that adverse infection events are frequent causes of death,(22–25) suggesting that safely targeting inflammation will be challenging. However, in our study, whilst we consistently observed that elevated CRP was associated with a greater relative risk of infection than cardiovascular death, absolute rates of infection death were still appreciably lower, suggesting that safer anti-inflammatory approaches could offer overall benefit. Indeed, the targeting of Rosuvastatin to people at high cardiovascular risk with CRP ≥2mg/L was shown to reduce inflammation, along with improving cardiovascular events *and* all-cause mortality.(19)

Beyond cardiovascular disease, our observations have much broader relevance since inflammation is causally implicated in many disease processes.(1–8) We observed that elevated CRP was common across diseases, and in disease- or multimorbidity-stratified analyses was almost uniformly associated with greater IRR for infection death than for cardiovascular death. Previously, we have shown that multimorbidity and some morbidities (class 3 obesity, hypertension, chronic respiratory disease, chronic kidney disease, psychiatric disease, chronic inflammatory and autoimmune rheumatological disease), along with advancing age and increasing SED, were associated with greater risk of infection death than other causes of death.(20) Since elevated CRP was still associated with greater risk of infection death than other causes of death in these subgroups, it is possible that the combination of elevated CRP and these diseases identifies people particularly predisposed to infection death.

An important question arising from our observations is which factors mediate the association between low-grade inflammation and infection death. Since elevated CRP was more strongly associated with infection death than cardiovascular or other causes of death in all but one of the morbidities we studied, a common mechanism (or mechanisms) seems the most plausible explanation. One possibility is that elevated CRP is a marker of frailty and reduced physiological reserve, and the substantial reduction in IRR between crude and age-sex adjusted models (**Appendix Table 7**) may support this possibility. However, the persistent association between elevated CRP and infection death in our ‘fully adjusted’ models suggests that factors beyond frailty and comorbidity are relevant. Another possibility is that elevated CRP is a biomarker of more broadly perturbed immune responses, as observed with ageing, and characterised by persistent low-grade inflammation and impaired adaptive immune responses to pathogens and vaccines.(26–28) Notably, recent data have suggested that clonal haematopoiesis of indeterminate potential, a disorder arising from somatic mutations that promote over-representation of pro-inflammatory myeloid clones (and elevated CRP),(29,30) is associated with increased risk of diverse infections.(31) Therefore, it will be important for future research to better profile the immune milieu in at risk populations, both in periods of usual health and during episodes of infection. These data may help to identify elements of the immune response whose perturbation may predispose to infection, which may act as a useful biomarker and even define safer avenues for anti-inflammatory therapy or strategies to reduce infection risk.

It is also important to interpret our work in the context of some limitations. First, the observational design precludes us from inferring causality in the association between low-grade systemic inflammation and infection death. Second, we have no data on the use of immunosuppressive anti-inflammatory therapies beyond the point of study enrolment, or indeed the lifetime doses of these. This is important since some people with undiagnosed inflammatory disorders may have later commenced these therapies, which are known to increase the risk of infection death. Moreover, lifetime dose of some immuno-suppressive agents is associated with increased risk of infection and cardiovascular events.(16,32) Hence, our adjusted IRR data may overestimate the association between elevated CRP and some causes of death. However, a relatively small proportion of the general population is prescribed such therapy, so our inability to account for incident use is unlikely to substantially diminish our estimates. Finally, it is important to note that CRP defines only one facet of inflammation and may not be the optimal biomarker to define or understand this issue; hence, future studies should explore other markers.

In conclusion, we show that elevated CRP is common in people with diverse chronic diseases and accumulates with multimorbidity. Irrespective of the threshold chosen, CRP defines a group of people at particularly increased relative risk of infection death. Moreover, this observation persisted in analyses restricted to the majority of comorbidities we studied, indicating that it is broadly relevant. This suggests that using CRP as a biomarker to identify people who may benefit from potent anti-inflammatory therapies also selects a population at increased risk of fatal infection, in keeping with recent clinical trial data in people with atherosclerosis.(9) This raises the question of whether more could be done to prevent infection, for example using enhanced vaccination strategies, before commencing anti-inflammatory therapy. Future research should aim to understand the immune responses to pathogens in people with low grade systemic inflammation, which may help to develop safer anti-inflammatory therapies for chronic disease and target their use to people most likely to obtain net benefit.

## Supporting information

STROBE

## Data Availability

All data produced in the present study are available upon application to UK Biobank (https://www.ukbiobank.ac.uk)

## Acknowledgements

This work was supported by the British Heart Foundation (FS/18/44/33792 and FS/12/80/29821). AWM was additionally supported by the NIHR Leeds Biomedical Research Centre and Diagnostic Evaluation Co-operative and a NIHR Senior Investigator award; the views expressed are those of the authors and not necessarily those of the National Health Service, the NIHR, or the Department of Health and Social Care.

