## Appendix Table for "Low grade systemic inflammation is a risk factor for infection death: an observational cohort study"

**Appendix Table 1: Self-reported disease category definitions**

| **NCD group** | **UK Biobank self-reported illnesses included** |
| --- | --- |
| Hypertension | Hypertension  Essential hypertension |
| Chronic cardiac disease | Angina  Cardiomyopathy  Heart attack/myocardial infarction  heart failure/pulmonary oedema  Hypertrophic cardiomyopathy  Coronary angioplasty (ptca) +/- stent  Coronary artery bypass grafts  Triple heart bypass |
| Chronic respiratory disease | Asthma  Bronchiectasis  Chronic obstructive airways disease/COPD  Emphysema  Emphysema/chronic bronchitis  Fibrosing alveolitis/unspecified alveolitis  Interstitial lung disease  Other chronic respiratory problems  Pulmonary fibrosis |
| Diabetes | Diabetes  Diabetic eye disease  Diabetic nephropathy  Diabetic neuropathy/ulcers  Type 1 diabetes  Type 2 diabetes |
| Cancer | Any cancer diagnosis during lifetime |
| Chronic liver disease | Liver failure/cirrhosis  Non-infective hepatitis  Oesophageal varices  Primary biliary cirrhosis |
| Chronic kidney disease | Diabetic nephropathy  Immunoglobulin A (IgA) nephropathy  Kidney nephropathy  Polycystic kidney  Renal failure not requiring dialysis  Renal failure requiring dialysis  Renal/kidney failure |
| Prior stroke/TIA | Brain haemorrhage  Ischaemic stroke  Stroke  Subarachnoid haemorrhage  Transient ischaemic attack (TIA) |
| Other neurology disease | Cerebral palsy  Epilepsy  Motor neurone disease  Multiple sclerosis  Myasthenia gravis  Parkinson’s disease |
| Psychiatric disease | Depression  Mania/bipolar disorder/manic depression  Postnatal depression  Schizophrenia |
| Chronic inflammatory and autoimmune rheumatic disease  (Rheumatological disease) | Ankylosing spondylitis  Dermatomyositis  Dermatopolymyositis  Giant cell/temporal arteritis  Myositis/myopathy  Polymyalgia rheumatica  Polymyositis  Psoriatic arthropathy  Rheumatoid arthritis  Sarcoidosis  Scleroderma/systemic sclerosis  Sjogren's syndrome/sicca syndrome  Systemic lupus erythematosus  Vasculitis |

All comorbidities are defined using self-reported illness at verbal nurse led interview (UK Biobank data fields 20002 and 20004). COPD, chronic obstructive pulmonary disease.

**Appendix Table 2: Non-steroidal anti-inflammatory and immuno-suppressive medication definitions**

| *Non-steroidal anti-inflammatory agents:* | |
| --- | --- |
| **Code** | **Name** |
| 1140925806 | aceclofenac |
| 1141163014 | acoflam 25mg e/c tablet |
| 1140928656 | advil 200mg tablet |
| 1140871514 | alrheumat 50mg capsule |
| 1141180152 | arcoxia 120mg tablet |
| 1141180148 | arcoxia 60mg tablet |
| 1141180150 | arcoxia 90mg tablet |
| 1140871320 | arthrofen 200 tablet |
| 1140871492 | arthrosin 250 tablet |
| 1140927086 | arthrotec 50 tablet |
| 1140871266 | arthrotec tablet |
| 1140871556 | arthroxen 250mg tablet |
| 1140871344 | artracin 25mg capsule |
| 1141192804 | bextra 10mg tablet |
| 1141192808 | bextra 20mg tablet |
| 1141192812 | bextra 40mg tablet |
| 1141169530 | brexidol 20mg tablet |
| 1140871374 | brufen 200mg tablet |
| 1140871660 | butacote 100mg e/c tablet |
| 1140853022 | butazolidin 100mg tablet |
| 1140871662 | butazone 100mg tablet |
| 1141191742 | calprofen 100mg/5ml s/f oral suspension |
| 1141176668 | celebrex 100mg capsule |
| 1141176670 | celebrex 200mg capsule |
| 1141176662 | celecoxib |
| 1140856312 | claradin 300mg tablet |
| 1140871606 | clinoril 100mg tablet |
| 1141176886 | closteril 100 m/r tablet |
| 1140928844 | clotam 200mg capsule |
| 1141163126 | condrotec tablet |
| 1140871388 | cuprofen 200mg tablet |
| 1141190952 | cuprofen plus tablet |
| 1141181872 | defanac 25mg e/c tablet |
| 1140851754 | defencin cp 40mg m/r capsule |
| 1141164746 | dexketoprofen |
| 1141162950 | dexomon sr 75mg m/r tablet |
| 1140884488 | diclofenac |
| 1140921828 | dicloflex 25mg e/c tablet |
| 1140917394 | diclomax sr 75mg m/r capsule |
| 1141146404 | diclotard 75 m/r tablet |
| 1141157112 | diclovol 25mg e/c tablet |
| 1140871260 | diclozip-25 e/c tablet |
| 1141153330 | difenor xl 100mg m/r tablet |
| 1140871282 | diflunisal |
| 1141145626 | digenac xl 100 m/r tablet |
| 1140871284 | dolobid 250mg tablet |
| 1140871886 | doloxene 65mg capsule |
| 1140868262 | doloxene compound capsule |
| 1140871450 | dysman-250 capsule |
| 1141173776 | ebretin 200mg capsule |
| 1140871386 | ebufac 200mg tablet |
| 1141193170 | eccoxolac 300mg capsule |
| 1141191028 | econac sr 75mg tablet |
| 1140871188 | etodolac |
| 1141180140 | etoricoxib |
| 1140871672 | feldene 10mg capsule |
| 1141188522 | feminax tablet |
| 1141182674 | fenactol 25mg e/c tablet |
| 1140917074 | fenoket 200mg m/r capsule |
| 1140871226 | fenoprofen |
| 1140871228 | fenopron 300mg tablet |
| 1141182868 | fenpaed 100mg/5ml s/f oral suspension |
| 1141181112 | feverfen 100mg/5ml s/f oral suspension |
| 1141152674 | flamatak mr 75mg m/r tablet |
| 1140871590 | flamatrol 10mg capsule |
| 1140871276 | flamrase 25mg e/c tablet |
| 1141170824 | flexin continus 25mg m/r tablet |
| 1140871360 | flexin-25 continus m/r tablet |
| 1141146400 | flexotard 100mg m/r tablet |
| 1141156966 | flexotard mr 100mg m/r tablet |
| 1140871236 | flurbiprofen |
| 1140871238 | froben 50mg tablet |
| 1141165574 | galprofen 100mg/5ml oral suspension |
| 1140853090 | ibular 200mg tablet |
| 1140853094 | ibumetin 200mg tablet |
| 1140871310 | ibuprofen |
| 1141157412 | ibuprofen product |
| 1140878030 | ibuprofen+codeine phosphate |
| 1141164050 | imazin xl 60mg/75mg m/r tablet |
| 1140871348 | imbrilon 25mg capsule |
| 1140871354 | indocid 25mg capsule |
| 1140853108 | indoflex 25mg capsule |
| 1140853110 | indolar 25mg capsule |
| 1140871442 | indomax 25 capsule |
| 1140909936 | indometacin |
| 1141181656 | indometacin product |
| 1140871434 | indomod 25mg m/r capsule |
| 1140925868 | inoven 200mg caplet |
| 1140871392 | isisfen 400mg tablet |
| 1141150922 | jomethid xl 200mg m/r capsule |
| 1140871404 | junifen 100mg/5ml s/f suspension |
| 1141164750 | keral 25mg tablet |
| 1140916866 | ketocid 200 m/r capsule |
| 1140871532 | ketonal 50mg capsule |
| 1140871506 | ketoprofen |
| 1140884558 | ketorolac |
| 1140927756 | ketotard 200 xl m/r capsule |
| 1140871528 | ketovail 100mg m/r capsule |
| 1140917406 | ketozip cr 200mg m/r capsule |
| 1141168652 | ketpron xl 100mg m/r capsule |
| 1140856314 | laboprin 300mg tablet |
| 1140863514 | laboprin dl 900mg sachet |
| 1140888872 | larafen cr 200mg m/r capsule |
| 1140871396 | lidifen 200mg tablet |
| 1140871196 | lodine 200mg tablet |
| 1140921968 | lofensaid 25 tablet |
| 1141181752 | mandafen 400mg tablet |
| 1140871542 | mefenamic acid |
| 1140916806 | meflam 250 capsule |
| 1140926732 | meloxicam |
| 1140877864 | migrafen 200mg tablet |
| 1140926796 | mobic 15mg tablet |
| 1140926794 | mobic 7.5mg tablet |
| 1140875350 | mobiflex 20mg tablet |
| 1140853012 | mobilan 25mg capsule |
| 1140909354 | motifene 75mg e/c+m/r capsule |
| 1140871406 | motrin 200mg tablet |
| 1140875336 | nabumetone |
| 1140871472 | naprosyn 250mg tablet |
| 1140871462 | naproxen |
| 1140881612 | naproxen+misoprostol |
| 1141187776 | nurofen 200mg tablet |
| 1140871568 | nycopren 250mg e/c tablet |
| 1140922318 | opustan 250mg capsule |
| 1141182642 | orbifen 100mg/5ml s/f oral suspension |
| 1140871516 | orudis 50mg capsule |
| 1140871522 | oruvail 100 m/r capsule |
| 1140877862 | pacifene 200mg tablet |
| 1141153346 | pardelprin mr 75mg m/r capsule |
| 1141180126 | parecoxib |
| 1140853100 | paxofen 200mg tablet |
| 1140856212 | paynocil 600mg tablet |
| 1140871654 | phenylbutazone product |
| 1140921974 | piroflam 10 capsule |
| 1140871666 | piroxicam |
| 1140871582 | pirozip 10 capsule |
| 1140871546 | ponstan 250mg capsule |
| 1140871490 | pranoxen continus 375mg m/r tablet |
| 1140925808 | preservex 100mg tablet |
| 1140877856 | proflex 200mg tablet |
| 1140868330 | progesic 200mg tablet |
| 1140871628 | prosaid 250mg tablet |
| 1140853082 | ramodar 200mg tablet |
| 1140877858 | relcofen 200mg tablet |
| 1140875338 | relifex 500mg tablet |
| 1140871482 | rheuflex-250 tablet |
| 1140911098 | rheumacin la 75mg m/r capsule |
| 1141167426 | rheumatac retard 75mg m/r tablet |
| 1140871180 | rhumalgan 25mg e/c tablet |
| 1140868294 | robaxisal forte tablet |
| 1141165244 | rofecoxib |
| 1140856438 | safapryn tablet |
| 1140856440 | safapryn-co tablet |
| 1140853018 | slo-indo 75mg m/r capsule |
| 1141145722 | slofenac sr 75mg m/r tablet |
| 1140928266 | solpaflex tablet |
| 1140856214 | solprin 300mg dispersible tablet |
| 1140871604 | sulindac |
| 1140871616 | surgam 200mg tablet |
| 1140868336 | synflex 275mg tablet |
| 1140875346 | tenoxicam |
| 1140871614 | tiaprofenic acid |
| 1141199380 | tiloket 50mg capsule |
| 1141183944 | tiloket cr 100mg m/r capsule |
| 1140864412 | timpron 250mg tablet |
| 1140875270 | tolectin 200mg capsule |
| 1140853030 | tolectin ds 400mg capsules |
| 1140928840 | tolfenamic acid |
| 1140875268 | tolmetin |
| 1140881118 | toradol 10mg tablet |
| 1141192798 | valdecoxib |
| 1141190644 | valdic 75 retard 75mg m/r tablet |
| 1140871256 | valenac 25mg e/c tablet |
| 1140871484 | valrox 250mg tablet |
| 1141165252 | vioxx 12.5mg tablet |
| 1141165312 | vioxx 12.5mg/5ml oral suspension |
| 1141165254 | vioxx 25mg tablet |
| 1141165314 | vioxx 25mg/5ml oral suspension |
| 1141184290 | vioxxacute 25mg tablet |
| 1141184292 | vioxxacute 50mg tablet |
| 1140871248 | volraman 25mg e/c tablet |
| 1140923920 | volsaid retard 75mg m/r tablet |
| 1140871168 | voltarol 25mg e/c tablet |

| *Immunosuppressive agents (including glucocorticoids):* | |
| --- | --- |
| **Code** | **Name** |
| 1140910598 | 5asa - mesalazine |
| 1141188588 | adalimumab |
| 1141180066 | anakinra |
| 1141166306 | arava 100mg tablet |
| 1141166302 | arava 10mg tablet |
| 1141166304 | arava 20mg tablet |
| 1140865580 | asacol 400mg e/c tablet |
| 1141188658 | asacol mr 400mg e/c tablet |
| 1140875400 | auranofin |
| 1140874484 | avloclor 250mg tablet |
| 1140869930 | azathioprine |
| 1141153242 | balsalazide disodium |
| 1140874790 | betamethasone |
| 1140874792 | betnelan 500mcg tablet |
| 1140874794 | betnesol 500mcg soluble tablet |
| 1140862572 | budesonide |
| 1141157418 | budesonide product |
| 1141163032 | calcort 1mg tablet |
| 1140874482 | chloroquine |
| 1140912244 | chloroquine 200mg tablet |
| 1140912246 | chloroquine 68mg/5ml syrup |
| 1141153248 | colazide 750mg capsule |
| 1141173346 | cortisone |
| 1140884704 | cortisone product |
| 1140874810 | cortistab 5mg tablet |
| 1140874814 | cortisyl 25mg tablet |
| 1140874104 | dapsone |
| 1140874822 | decadron 500micrograms tablet |
| 1140868370 | decortisyl 5mg tablet |
| 1141145782 | deflazacort |
| 1140874936 | deltacortril enteric 2.5mg e/c tablet |
| 1140910634 | deltahydrocortisone |
| 1140874940 | deltastab 1mg tablet |
| 1140874816 | dexamethasone |
| 1141176842 | dexsol 2mg/5ml oral solution |
| 1140865742 | dipentum 250mg capsule |
| 1140926004 | entocort cr 3mg m/r capsule |
| 1140874784 | florinef 100mcg tablet |
| 1140884672 | fludrocortisone |
| 1140910424 | hc - hydrocortisone |
| 1141188594 | humira 40mg injection solution 0.8ml prefilled syringe |
| 1140874896 | hydrocortisone |
| 1141157294 | hydrocortisone product |
| 1140874954 | hydrocortistab 20mg tablet |
| 1140874956 | hydrocortone 10mg tablet |
| 1140884308 | hydroxychloroquine |
| 1141145996 | imuran 10mg tablet |
| 1141180070 | kineret 100mg/0.67ml injection prefilled syringe |
| 1140874978 | medrone 2mg tablet |
| 1140865578 | mesalazine |
| 1141189390 | mesren mr 400mg m/r tablet |
| 1140869848 | methotrexate |
| 1140874976 | methylprednisolone |
| 1140910036 | mtx - methotrexate |
| 1140879502 | olsalazine |
| 1140857534 | oradexon 500micrograms tablet |
| 1141156738 | paludrine tablet+avloclor tablet 100mg/250mg travel pack |
| 1140875316 | penicillamine |
| 1140865588 | pentasa sr 250mg m/r tablet |
| 1140875392 | plaquenil 200mg tablet |
| 1140874944 | precortisyl 1mg tablet |
| 1140874950 | prednesol 5mg tablet |
| 1140874930 | prednisolone |
| 1141157402 | prednisolone product |
| 1140868364 | prednisone |
| 1141156676 | proguanil hydrochloride+chloroquine phosphate |
| 1140875404 | ridaura 3mg tablet |
| 1141188196 | ridaura tiltab 3mg tablet |
| 1140865670 | salazopyrin 500mg tablet |
| 1140865598 | salofalk 250mg e/c tablet |
| 1140857672 | sintisone 5mg tablet |
| 1141188900 | sulazine ec 500mg e/c tablet |
| 1140909702 | sulfasalazine |
| 1140865668 | sulphasalazine |
| 1140868426 | triamcinolone |

**Appendix Table 3: Cardiovascular, infection and other causes of death definitions**

**Cardiovascular death:**

I00-I99, excluding infection codes: [I32.0, I32.1, I33.0, I33.9, I38, I39, I40.0, I41.0-I41.2, I43.0, I52.0-I52.1, I68.1, I98.1]

**Infection Death:**

A00.0, A00.1, A00.9, A01.0-A01.4, A02.0-A02.2, A02.8, A02.9, A03.0-A03.3, A03.8, A03.9, A04.0-A04.9, A06.2, A06.4-A06.6, A07.0, A07.8, A07.9, A08.0-A08.4, A09.9, A15.0-A15.9, A16.0-A16.9, A17.0, A17.1, A17.8, A17.9, A18.0-A18.8, A19.0-A19.2, A19.8, A19.9, A20.0-A20.3, A20.7-A20.9, A21.0-A21.3, A21.7-A21.9, A22.0-A22.2, A22.7-A22.9, A23.0-A23.3, A23.8, A23.9, A24.0-A24.4, A25.0, A25.9, A26.7-A26.9, A27.0, A27.8, A27.9, A28.8, A28.9, A30.0-A30.5, A30.8, A30.9, A31.0, A31.1, A31.8, A31.9, A32.0, A32.1, A32.7-A32.9, A33.X, A34.X, A35.X, A36.0-A36.3, A36.8, A36.9, A37.0, A37.1, A37.8, A37.9, A39.0, A39.2-A39.5, A39.8, A39.9, A40.0-A40.3, A40.8, A40.9, A41.0, A41.0-A41.5, A41.8, A41.9, A42.0-A42.2, A42.7-A42.9, A46.X, A48.0, A48.8, A49.0-A49.3, A49.2, A49.8, A49.9, A54.0, A54.2-A54.4, A54.5, A54.6, A54.8, A54.9, A56.4, A57.X, A66.0-A66.4, A66.6-A66.9, A67.0-A67.3, A67.9, A68.0, A68.1, A68.9, A69.1, A70.X, A71.0, A71.1, A71.9, A74.0, A74.8, A74.9, A75.0-A75.3, A75.9, A77.0, A79.0, A79.8, A79.9, A80.9, A81.0, A81.8, A81.9, A82.0, A82.1, A82.9, A83.0, A83.6, A83.8, A83.9, A84.0, A84.8, A84.9, A85.0-A85.2, A85.8, A86.X, A87.0, A87.1, A87.2, A87.8, A87.9, A88.0, A88.8, A89.X, A90.X, A91.X, A92.0, A92.3, A92.8, A92.9, A93.0, A93.8, A94.X, A95.0, A95.1, A95.9, A96.0, A96.8, A96.9, A98.0, A98.3, A98.4, A98.8, A99.X, B00.0, B00.1, B00.2, B00.3-B00.5, B00.7-B00.9, B01.0, B01.1, B01.2, B01.8, B01.9, B02.0, B02.1, B02.2, B02.3, B02.7-B02.9, B03.X, B05.0, B05.1, B05.2, B05.3, B05.4, B05.8, B05.9, B06.0, B06.8, B06.9, B07.X, B08.0, B08.4, B08.5, B08.8, B09.X, B16.0-B16.2, B16.9, B17.0, B17.1, B17.8, B17.9, B18.0-B18.2, B18.8, B18.9, B19.0, B19.9, B25.0, B25.1, B25.2, B25.8, B25.9, B26.0, B26.1, B26.2, B26.3, B26.8, B26.9, B27.0, B27.1, B27.8, B27.9, B30.0-B30.2, B30.3, B30.8, B30.9, B33.0, B33.2-B33.4, B33.8, B34.0-B34.4 (exclude corona), B34.8, B34.9, B35.9, B36.8, B37.0-B37.9, B38.0-B38.4, B38.7-B38.9, B41.0, B41.7-B41.9, B43.1, B43.2, B44.0-B44.2, B44.7-B44.9, B45.0-B45.3, B45.7-B45.9, B46.8, B47.1, B48.7, B48.8, B50.0, B50.8, B50.9, B51.0, B51.8, B51.9, B52.0, B52.8, B52.9, B53.0, B53.1, B53.8, B54.X, B55.0-B55.2, B55.9, B56.0, B56.1, B56.9, B58.0-B58.3, B58.8, B58.9, B60.0, B60.8, B64.X, B65.0-B65.2, B65.8, B65.9, B67.8, B67.9, B71.0, B71.8, B71.9, B76.1, B81.0, B81.4, B81.8, B82.0, B82.9, B83.0, B83.8, B83.9, B85.0-B85.2, B85.4, B87.0-B87.4, B87.8, B87.9, B88.0, B89.X, B91.X, B92.X, B94.0, B94.0, B94.1, B94.2, B94.8, B94.9, B95.0-B95.8, B96.0-B96.2, B96.3, B96.4, B96.5, B96.7, B96.8, B97.0-B97.8, D47.5, D73.3, E32.1, G00.0-G00.3, G00.8, G00.9, G01.X, G02.0, G02.1, G02.8, G03.0-G03.2, G03.8, G03.9, G04.2, G05.0, G05.1, G05.2, G06.0-G06.2, G07X, G14.X, G53.0, G53.1, G63.0, G73.1, G73.4, G93.3, G94.0, H03.0, H06.1, H10.0, H10.2, H10.3, H10.4, H10.5, H10.8, H10.9, H13.0, H13.1, H13.2, H16.1, H16.2, H19.1, H19.2, H19.3, H22.0, H32.0, H60.0, H60.1, H60.2, H60.3, H60.5, H60.8, H60.9, H62.0, H62.1, H62.3, H62.4, H65.0, H65.1, H65.2-H65.4, H65.9, H66.0-H66.4, H66.9, H67.0, H67.1, H67.8, H70.0, H70.1, H70.8, H70.9, H73.0, H73.1, H75.0, H94.0, I32.0, I32.1, I41.0, I41.1, I41.2, I42.3, I43.0, I52.0, I52.1, I68.1, I98.1, J00.X, J01.0-J01.4, J01.8, J01.9, J02.0, J02.8, J02.9, J03.0, J03.0, J03.8, J03.9, J06.0, J06.8, J06.9, J09.X, J10.0, J10.0, J10.1, J10.8, J11.0, J11.1, J11.8, J12.0, J12.1, J12.2, J12.3, J12.8, J12.9, J13X, J14X, J15.0- J15.9, J16.0, J16.8, J17.0, J17.1, J17.2, J17.3, J17.8, J18.0-J18.2, J18.8, J18.9, J20.0, J20.1, J20.2, J20.3, J20.4, J20.5-J20.7, J21.0, J21.1, J22.X, J31.0-J31.2, J32.0-J32.4, J32.8, J32.9, J34.0, J35.0, J36X, J39.0, J39.1, J44.0, J65X, J85.0, J85.1, J85.2, J85.3, K04.6, K04.7, K11.3, K12.2, K23.0, K35.2, K35.3, K35.8, K36.X, K37.X, K51.5, K52. 3, K57.0, K57.1, K57.2, K57.3, K57.4, K57.5, K57.8, K57.9, K61.0-K61.4, K63.0, K65.0, K65.8, K65.9, K67.0-K67.3, K75.0, K77.0, K80.0-K80.5, K81.0, K81.1, K81.9, K83.0, K93.0, L00.X, L02.0-L02.4, L02.8, L02.9, L03.0-L03.3, L03.8, L03.9, L05.0, L05.9, L92.2, L98.3, M00.0-M00.2, M00.8, M01, M01.0, M01.1, M01.3, M01.4, M01.5, M01.6, M02.3, M03.0, M03.1, M35.4, M46.2, M49.0-M49.2, M49.3, M63.0, M63.1, M65.0, M68.0, M71.0, M73.0, M86.0-M86.2, M86.3-M86.6, M86.8, M86.9, M89.6, N08.0, N15.1, N16.0, N22.0, N29.1, N33.0, N34.0, N35.1, N39.0, N41.2, N45.0, N45.9, N73.0, N73.1, N73.2, N73.3, N73.4, N73.5, N74.0, N74.1, N75.1, N76.4, N77.0, N77.1, O26.4, O35.3, O75.3, O98.0, O98.4, O98.5, O98.6, O98.8, O98.9, R57.2

**Other**

All other deaths not categorised as cardiovascular or infection

**Appendix Table 4: Characteristics of UK Biobank participants with missing CRP data**

|  | **CRP data available**  **(461,052)** | **No CRP data available**  **(32,200)** |
| --- | --- | --- |
| **Age** |  |  |
| **<45** | 47330 (10.3) | 3364 (10.5) |
| **45 to <50** | 60575 (13.1) | 4276 (13.3) |
| **50 to <55** | 70021 (15.2) | 4918 (15.3) |
| **55 to <60** | 83410 (18.1) | 5845 (18.2) |
| **60 to <65** | 111734 (24.2) | 7631 (23.7) |
| **65+** | 87982 (19.1) | 6166 (19.2) |
| **Sex** |  |  |
| **Female** | 250435 (54.3) | 18395 (57.1) |
| **Male** | 210617 (45.7) | 13805 (42.9) |
| **Ethnicity** |  |  |
| **White** | 437071 (94.8) | 30091 (93.5) |
| **Ethnic minority** | 23981 (5.2) | 2109 (6.6) |
| **SED quintile** |  |  |
| **1 (least deprived)** | 92615 (20.1) | 6074 (18.9) |
| **2** | 92404 (20.0) | 6209 (19.3) |
| **3** | 92312 (20.0) | 6341 (19.7) |
| **4** | 92262 (20.0) | 6387 (19.8) |
| **5 (most deprived)** | 91459 (19.8) | 7189 (22.3) |
| **Smoking** |  |  |
| **Never** | 252445 (54.8) | 17763 (55.2) |
| **Former** | 160153 (34.7) | 10849 (33.7) |
| **Current** | 48454 (10.5) | 3588 (11.1) |
| **Obesity** |  |  |
| **Not obese** | 347843 (75.5) | 23631 (73.4) |
| **Class 1** | 81342 (17.6) | 5867 (18.2) |
| **Class 2** | 23015 (5.0) | 1901 (5.9) |
| **Class 3** | 8852 (1.9) | 801 (2.5) |
| **Hypertension** | 122101 (26.5) | 8689 (27) |
| **Chronic cardiac disease** | 21797 (4.7) | 1668 (5.2) |
| **Chronic respiratory disease** | 59587 (12.9) | 4312 (13.4) |
| **Diabetes** | 22992 (5.0) | 1758 (5.5) |
| **Cancer** | 37821 (8.2) | 3185 (9.9) |
| **Chronic liver disease** | 903 (0.2) | 53 (0.2) |
| **Chronic kidney disease** | 1175 (0.3) | 96 (0.3) |
| **Prior stroke/TIA** | 7979 (1.7) | 613 (1.9) |
| **Other neurological disease** | 6088 (1.3) | 458 (1.4) |
| **Psychiatric disorder** | 27759 (6.0) | 1886 (5.9) |
| **Rheumatological disease** | 10395 (2.3) | 697 (2.2) |

Legend: UKB recruited 502,505 participants and CRP data is available for 468,528 of these. Missing data for comorbidities (n=863), body mass index (n=3,106), smoking (n=2,949), ethnicity (n=2,777), and SED (n=624), and loss to follow‐up or withdrawal of consent (n=1,343), resulted in exclusion of 7,476 participants from our analyses (some participants with more than one variable missing), resulting in a study cohort of 461,052 participants with available CRP data and 32,200 participants without CRP data for whom we have all of the other phenotyping data to present in this table. CRP – C-reactive protein; SED – socio-economic deprivation; TIA – transient ischaemic attack.

**Appendix Table 5: Characteristics of participants with CRP ≥5mg/L**

|  | **CRP<5**  n=407,584 | **CRP ≥5**  n=53,468 |
| --- | --- | --- |
| **Age** |  |  |
| **<45** | 42,792 (10.5) | 4,538 (8.5) |
| **45 to <50** | 54,585 (13.4) | 5,990 (11.2) |
| **50 to <55** | 62,259 (15.3) | 7,762 (14.5) |
| **55 to <60** | 73,869 (18.1) | 9,541 (17.8) |
| **60 to <65** | 97,968 (24) | 13,766 (25.7) |
| **65+** | 76,111 (18.7) | 11,871 (22.2) |
| **Sex** |  |  |
| **Female** | 217,953 (53.5) | 32,482 (60.8) |
| **Male** | 189,631 (46.5) | 20,986 (39.2) |
| **Ethnicity** |  |  |
| **White** | 386,705 (94.9) | 50,366 (94.2) |
| **Non-white** | 20,879 (5.1) | 3,102 (5.8) |
| **SED quintile** |  |  |
| **1 (least deprived)** | 83,400 (20.5) | 8,889 (16.6) |
| **2** | 82,828 (20.3) | 9,306 (17.4) |
| **3** | 82,260 (20.2) | 9,959 (18.6) |
| **4** | 81,111 (19.9) | 11,090 (20.7) |
| **5 (most deprived)** | 77,985 (19.1) | 14,224 (26.6) |
| **Smoking** |  |  |
| **Never** | 226,697 (55.6) | 25,748 (48.2) |
| **Former** | 140,683 (34.5) | 19,470 (36.4) |
| **Current** | 40,204 (9.9) | 8,250 (15.4) |
| **Obesity** |  |  |
| **Not obese** | 320,999 (78.8) | 26,844 (50.2) |
| **Class 1** | 66,687 (16.4) | 14,655 (27.4) |
| **Class 2** | 15,665 (3.8) | 7,350 (13.7) |
| **Class 3** | 4,233 (1) | 4,619 (8.6) |
| **Hypertension** | 102,357 (25.1) | 19,744 (36.9) |
| **Chronic cardiac disease** | 18,457 (4.5) | 3,340 (6.2) |
| **Chronic respiratory disease** | 49,662 (12.2) | 9,925 (18.6) |
| **Diabetes** | 18,818 (4.6) | 4,174 (7.8) |
| **Cancer** | 32,411 (8) | 5,410 (10.1) |
| **Chronic liver disease** | 720 (0.2) | 183 (0.3) |
| **Chronic kidney disease** | 930 (0.2) | 245 (0.5) |
| **Prior stroke/TIA** | 6,651 (1.6) | 1,328 (2.5) |
| **Other neurological disease** | 5,050 (1.2) | 1,038 (1.9) |
| **Psychiatric disorder** | 23,134 (5.7) | 4,625 (8.7) |
| **Rheumatological disease** | 7,499 (1.8) | 2,896 (5.4) |
| **NSAID use** | 63,966 (15.6) | 11,207 (21.0) |
| **Immunosuppressant use** | 9,097 (2.2) | 3,084 (5.8) |
| **Number of chronic diseases** |  |  |
| **0** | 183,751 (45.1) | 11,660 (21.8) |
| **1** | 133,589 (32.8) | 18,051 (33.8) |
| **2** | 61,408 (15.1) | 13,928 (26.0) |
| **3+** | 28,836 (7.1) | 9,829 (18.4) |

Legend: CRP – C-reactive protein; NSAID – non-steroidal anti-inflammatory drug; SED – socio-economic deprivation; TIA – transient ischaemic attack.

**Appendix Table 6: Characteristics of participants with CRP ≥10mg/L**

|  | **CRP<10**  n=442,028 | **CRP ≥10**  n=19,024 |
| --- | --- | --- |
| **Age** |  |  |
| **<45** | 45,702 (10.3) | 1,628 (8.6) |
| **45 to <50** | 58,506 (13.2) | 2,069 (10.9) |
| **50 to <55** | 67,290 (15.2) | 2,731 (14.4) |
| **55 to <60** | 80,060 (18.1) | 3,350 (17.6) |
| **60 to <65** | 106,814 (24.2) | 4,920 (25.9) |
| **65+** | 83,656 (18.9) | 4,326 (22.7) |
| **Sex** |  |  |
| **Female** | 239,142 (54.1) | 11,293 (59.4) |
| **Male** | 202,886 (45.9) | 7,731 (40.6) |
| **Ethnicity** |  |  |
| **White** | 419,125 (94.8) | 17,946 (94.3) |
| **Non-white** | 22,903 (5.2) | 1,078 (5.7) |
| **SED quintile** |  |  |
| **1 (least deprived)** | 89,217 (20.2) | 3,072 (16.1) |
| **2** | 88,842 (20.1) | 3,292 (17.3) |
| **3** | 88,723 (20.1) | 3,496 (18.4) |
| **4** | 88,282 (20) | 3,919 (20.6) |
| **5 (most deprived)** | 86,964 (19.7) | 5,245 (27.6) |
| **Smoking** |  |  |
| **Never** | 243,451 (55.1) | 8,994 (47.3) |
| **Former** | 153,080 (34.6) | 7,073 (37.2) |
| **Current** | 45,497 (10.3) | 2,957 (15.5) |
| **Obesity** |  |  |
| **Not obese** | 337,916 (76.4) | 9,927 (52.2) |
| **Class 1** | 76,834 (17.4) | 4,508 (23.7) |
| **Class 2** | 20,552 (4.6) | 2,463 (12.9) |
| **Class 3** | 6,726 (1.5) | 2,126 (11.2) |
| **Hypertension** | 114,902 (26) | 7,199 (37.8) |
| **Chronic cardiac disease** | 20,509 (4.6) | 1,288 (6.8) |
| **Chronic respiratory disease** | 55,726 (12.6) | 3,861 (20.3) |
| **Diabetes** | 21,367 (4.8) | 1,625 (8.5) |
| **Cancer** | 35,757 (8.1) | 2,064 (10.8) |
| **Chronic liver disease** | 826 (0.2) | 77 (0.4) |
| **Chronic kidney disease** | 1,061 (0.2) | 114 (0.6) |
| **Prior stroke/TIA** | 7,476 (1.7) | 503 (2.6) |
| **Other neurological disease** | 5,653 (1.3) | 435 (2.3) |
| **Psychiatric disorder** | 26,049 (5.9) | 1,710 (9) |
| **Rheumatological disease** | 9,001 (2) | 1,394 (7.3) |
| **NSAID use** | 70,886 (16.0) | 4,287 (22.5) |
| **Immunosuppressant use** | 10,709 (2.4) | 1,472 (7.7) |
| **Number of chronic diseases** |  |  |
| **0** | 191,331 (43.3) | 4,080 (21.4) |
| **1** | 145,499 (32.9) | 6,141 (32.3) |
| **2** | 70,351 (15.9) | 4,985 (26.2) |
| **3+** | 34,847 (7.9) | 3,818 (20.1) |

Legend: CRP – C-reactive protein; NSAID – non-steroidal anti-inflammatory drug; SED – socio-economic deprivation; TIA – transient ischaemic attack.

**Appendix Table 7: Association between CRP ≥2mg/L and cause-specific mortality using partially and fully adjusted models**

|  | **IRR (95% CI)** | | |
| --- | --- | --- | --- |
| **Model** | **CV death** | **Infection death** | **Other death** |
| **Age + sex** |  |  |  |
| CRP ≥2 mg/L | 1.95 (1.85-2.06) | 2.33 (2.09-2.61) | 1.62 (1.58-1.67) |
| **As above + socio-demographic factors + co-morbidity** | | | |
| CRP ≥2 mg/L | 1.48 (1.40-1.57) | 1.72 (1.53-1.93) | 1.41 (1.37-1.45) |
| **As above + NSAID use** | | | |
| CRP ≥2 mg/L | 1.48 (1.35-1.77) | 1.71 (1.52-1.93) | 1.41 (1.37-1.45) |
| **As above + immuno-suppressant use (‘fully adjusted’)** | | | |
| CRP ≥2 mg/L | 1.48 (1.40-1.57) | 1.70 (1.51-1.92) | 1.41 (1.37-1.45) |

Legend: CI – confidence interval; CRP – C-reactive protein; CV – cardiovascular; IRR – incidence rate ratio; NSAID – non-steroidal anti-inflammatory drug.

**Appendix Table 8: Association between CRP ≥2 mg/L, CRP ≥5 mg/L or CRP ≥10mg/L and cause-specific mortality using fully adjusted models after exclusion of participants who died during the first 6-months of follow-up**

|  | **IRR (95% CI)** | | |
| --- | --- | --- | --- |
|  | **CV death** | **Infection death** | **Other death** |
| CRP ≥2 mg/L | 1.47 (1.39-1.56) | 1.70 (1.51-1.91) | 1.39 (1.35-1.43) |
| CRP ≥5 mg/L | 1.55 (1.44-1.66) | 1.91 (1.67-2.17) | 1.51 (1.45-1.56) |
| CRP ≥10 mg/L | 1.58 (1.43-1.74) | 2.16 (1.82-2.55) | 1.70 (1.62-1.80) |

Legend: CI – confidence interval; CRP – C-reactive protein; CV – cardiovascular; IRR – incidence rate ratio.

**Appendix Table 9: Association between CRP ≥5 mg/L and cause-specific mortality stratified by individual comorbidity groups**

|  | **IRR (95% CI)** | | |
| --- | --- | --- | --- |
| **Disease** | **CV death** | **Infection death** | **Other death** |
| **Obesity Class 1** | 1.44 (1.26-1.65) | 1.93 (1.47 - 2.53) | 1.51 (1.40-1.62) |
| **Obesity Class 2** | 1.29 (1.06-1.57) | 1.70 (1.10 - 2.63) | 1.17 (1.04-1.32) |
| **Obesity Class 3** | 1.36 (1.05-1.76) | 2.51 (1.46 - 4.31) | 1.08 (0.90-1.29) |
| **Hypertension** | 1.55 (1.41-1.70) | 1.71 (1.43 - 2.06) | 1.47 (1.39-1.56) |
| **Chronic cardiac disease** | 1.61 (1.41-1.84) | 2.02 (1.49 - 2.73) | 1.50 (1.34-1.67) |
| **Chronic respiratory disease** | 1.42 (1.23-1.65) | 2.27 (1.81 - 2.84) | 1.58 (1.46-1.71) |
| **Diabetes** | 1.80 (1.56-2.09) | 1.73 (1.26 - 2.36) | 1.31 (1.17-1.46) |
| **Cancer** | 1.44 (1.16-1.79) | 2.02 (1.43 - 2.87) | 1.82 (1.68-1.96) |
| **Chronic kidney disease** | 2.18 (1.29-3.68) | 5.71 (2.49 - 13.07) | 1.78 (1.24-2.56) |
| **Prior stroke/TIA** | 1.35 (1.06-1.71) | 1.86 (1.16 - 2.98) | 1.62 (1.37-1.92) |
| **Other neurological disease** | 1.57 (1.06-2.32) | 1.18 (0.58 - 2.38) | 1.29 (1.04-1.59) |
| **Psychiatric disorder** | 1.49 (1.18-1.87) | 2.06 (1.41 - 3.01) | 1.55 (1.37-1.77) |
| **Rheumatological disease** | 1.51 (1.15-1.99) | 1.79 (1.16 - 2.75) | 1.59 (1.35-1.86) |

Legend: IRRs denote risk of cause-specific death associated with CRP ≥5 mg/L versus CRP <5 mg/L in disease strata. CI – confidence interval; CRP – C-reactive protein; CV – cardiovascular; IRR – incidence rate ratio; TIA – transient ischaemic attack.

**Appendix Table 10: Association between CRP ≥10mg/L and cause-specific mortality stratified by individual comorbidity groups**

|  | **IRR (95% CI)** | | |
| --- | --- | --- | --- |
| **Disease** | **CV death** | **Infection death** | **Other death** |
| **Obesity Class 1** | 1.59 (1.30 - 1.95) | 2.36 (1.64 - 3.39) | 1.83 (1.65 - 2.03) |
| **Obesity Class 2** | 1.54 (1.19 - 2.01) | 1.73 (0.98 - 3.06) | 1.31 (1.11 - 1.55) |
| **Obesity Class 3** | 1.42 (1.07 - 1.89) | 2.05 (1.25 - 3.37) | 1.16 (0.95 - 1.43) |
| **Hypertension** | 1.58 (1.39 - 1.80) | 1.98 (1.56 - 2.50) | 1.67 (1.55 - 1.81) |
| **Chronic cardiac disease** | 1.60 (1.33 - 1.92) | 1.81 (1.20 - 2.73) | 1.71 (1.48 - 1.99) |
| **Chronic respiratory disease** | 1.37 (1.12 - 1.67) | 2.45 (1.87 - 3.21) | 1.71 (1.54 - 1.91) |
| **Diabetes** | 1.71 (1.40 - 2.09) | 1.84 (1.23 - 2.75) | 1.42 (1.23 - 1.65) |
| **Cancer** | 1.31 (0.96 - 1.79) | 2.13 (1.37 - 3.34) | 2.27 (2.05 - 2.50) |
| **Chronic kidney disease** | 1.48 (0.72 - 3.06) | 10.22 (4.32 - 24.19) | 2.30 (1.51 - 3.52) |
| **Prior stroke/TIA** | 1.17 (0.82 - 1.66) | 3.23 (1.88 - 5.54) | 1.82 (1.46 - 2.27) |
| **Other neurological disease** | 1.14 (0.64 - 2.02) | 1.47 (0.60 - 3.58) | 1.60 (1.22 - 2.10) |
| **Psychiatric disorder** | 1.53 (1.11 - 2.11) | 1.88 (1.15 - 3.07) | 1.77 (1.49 - 2.10) |
| **Rheumatological disease** | 1.79 (1.31 - 2.45) | 2.48 (1.56 - 3.95) | 1.45 (1.20 - 1.76) |

Legend: IRRs denote risk of cause-specific death associated with CRP ≥10 mg/L versus CRP <10 mg/L in disease strata. CI – confidence interval; CRP – C-reactive protein; CV – cardiovascular; IRR – incidence rate ratio; TIA – transient ischaemic attack.

**Appendix Table 11: Association between CRP ≥5/L and cause-specific mortality stratified by number of morbidities**

| **Number of comorbidities** | **IRR (95% CI)** | | |
| --- | --- | --- | --- |
|  | **CV death** | **Infection death** | **Other death** |
| **0** | 1.73 (1.42-2.11) | 1.85 (1.26-2.72) | 1.53 (1.40-1.67) |
| **1** | 1.54 (1.34-1.77) | 1.85 (1.43-2.40) | 1.53 (1.43-1.63) |
| **2** | 1.54 (1.36-1.74) | 1.93 (1.53-2.43) | 1.47 (1.37-1.57) |
| **3+** | 1.48 (1.32-1.65) | 1.89 (1.53-2.34) | 1.42 (1.32-1.53) |

Legend: IRRs denote risk of cause-specific death associated with CRP ≥5 mg/L versus CRP <5 mg/L in multimorbidity strata. CI – confidence interval; CRP – C-reactive protein; CV – cardiovascular; IRR – incidence rate ratio; TIA – transient ischaemic attack.

**Appendix Table 12: Association between CRP ≥10/L and cause-specific mortality stratified by number of morbidities**

| **Number of comorbidities** | **IRR (95% CI)** | | |
| --- | --- | --- | --- |
|  | **CV death** | **Infection death** | **Other death** |
| **0** | 1.90 (1.43-2.54) | 2.19 (1.27-3.78) | 1.76 (1.54-2.01) |
| **1** | 1.78 (1.46-2.18) | 1.87 (1.28-2.75) | 1.85 (1.68-2.03) |
| **2** | 1.49 (1.24-1.80) | 3.10 (2.35-4.08) | 1.68 (1.52-1.85) |
| **3+** | 1.58 (1.36-1.84) | 2.04 (1.56-2.67) | 1.72 (1.57-1.89) |

Legend: IRRs denote risk of cause-specific death associated with CRP ≥10 mg/L versus CRP <10 mg/L in multimorbidity strata. CI – confidence interval; CRP – C-reactive protein; CV – cardiovascular; IRR – incidence rate ratio; TIA – transient ischaemic attack.
